# Increasing trends in mental health problems among Chinese young adolescents: results from repeated cross-sectional data in Changsha 2016-2020

**DOI:** 10.1101/2021.12.04.21267298

**Authors:** Zhipeng Wu, Biao Wang, Zhulin Zou, Zhening Liu, Xudong Chen, Yicheng Long

## Abstract

This study performed a repeated cross-sectional analysis to explore possible trends in mental health problems among Chinese adolescents during 2016 to 2020. A total of 2837 seventh-grade students from Changsha city were surveyed in three waves in 2016, 2019 and 2020. The results showed that obsessive-compulsive tendencies, interpersonal sensitivity, depression, anxiety, academic stress and emotional disturbance problems were significantly increased in surveyed adolescents during 2016 to 2020. Moreover, positive rates of most of these problems were higher in females than males, and were significantly increased in only females. These results highlight the importance of focusing on mental health among Chinese adolescents, especially girls.

## 1. Introduction

Childhood and early adolescence are crucial periods for mental health development with particularly high risk for mental health problems (Garcia-Carrion et al., 2019; Kessler et al., 2005). These problems can persist into adulthood and even lead to serious mental disease if they are undetected or not treated appropriately (Pine et al., 1998; Zeng et al., 2019; Zhang et al., 2019).

Over the past decades, China has witnessed great developments in economy and society; however, multiple studies have reported a dramatic decrease of mental health in China, especially in Chinese adolescents during the same period (Feng et al., 2020; Su and Liu, 2020; Wang and Tapia Granados, 2019; Xin et al., 2020). For example, a recent cross-temporal meta-analysis showed that Chinese adolescents’ depression was significantly increased from 1989 to 2018 (Su and Liu, 2020). Another cross-temporal meta-analysis, which was based on data between 1992 and 2017, also reported that Chinese adolescents’ anxiety level was significantly increased since 1992 (Xin et al., 2020). To improve public mental health, it is thus important to assess the prevalence and changes over time in prevalence of mental health problems in Chinese adolescents (Su and Liu, 2020).

Although having gained important insights into trends in mental health problems in Chinese adolescents in recent years, the above-mentioned published literatures are limited in several ways. First, reports on newly trends after the year 2018 are sparse. Second, most of them only focused on a single dimension of mental health such as depression or anxiety, while a more comprehensive survey in multiple dimensions is rare. Third, most of these studies are cross-temporal meta-analyses, which pooled previous studies using the same mental health assessing instruments (e.g., Self-rating Depression Scale (Su and Liu, 2020)) together. However, such an approach may not be able to fully address the potential bias caused by regional diversity (e.g., the pooled studies were across different provinces in China with different economic and educational levels (Su and Liu, 2020)).

In this study, we performed a repeated cross-sectional analysis to overcome the above limitations. Repeated cross-sectional studies utilize the same instruments and methodologies in the same population over multiple time timepoints; therefore, they were thought to be most suitable for investigating trends of mental health statuses over time (Myhr et al., 2020; Richter et al., 2019). In specific, a total of 2837 adolescents from a sample high school in Changsha city, China, were surveyed in three waves from 2016 to 2020. Mental health status was assessed using a validated multi-dimensional mental health scale, Mental Health Inventory of Middle School Students (MMHI-60) (Wang et al., 1997; Wu et al., 2021a) and compared between different years.

## 2. Methods

This study uses data collected from 2016 to 2020 in a total of 2837 Chinese junior high school students in three waves (978 students in September 2016; 949 students in September 2019; and 910 students in September 2020). All participants were new seventh-grade students from the same campus of a high school (Tianding campus of The High School Attached to Hunan Normal University and Bocai Experimental School in Changsha city, Hunan province, China) when they completed the survey. Note that participants with missing data for any item of the MMHI-60 in the surveys have been excluded from the study. All participants and their supervisors provided written informed consents and the study was approved by the Ethics Committees of the Second Xiangya Hospital of Central South University, Changsha.

All participants completed the MMHI-60 in the classroom to assess their mental health statuses. MMHI-60 is a validated and widely used self-report measure of mental health problems, which includes 10 subscales of distinct dimensions (60 items in total, and 6 items for each subscale): obsessive–compulsive tendencies, paranoid ideation, hostility, interpersonal sensitivity, depression, anxiety, academic stress, maladaptation, emotional disturbance and psychological imbalance (Du et al., 2019; Luo et al., 2020; Wang et al., 1997; Wu et al., 2021a; Xiong et al., 2021). All items were scored from 1 to 5, and higher scores indicate more severe mental health problems. According to Wang et al. (Wang et al., 1997), a cutoff of average points of a subscale >=2 is identified as having a mental health problem (positive for that subscale). This cutoff has shown good specificity and sensitivity in previous research (Du et al., 2019; Luo et al., 2020; Zhao and Liao, 2016). Positive rates of each subscale were calculated based on such a cutoff, and Cochran–Armitage trend tests were used to determine if there are increasing or decreasing trends of positive rates during the period of 2016-2020 (Yang et al., 2017). Moreover, considering that sex differences in mental health among adolescents have been widely reported (Kim et al., 2018; Luo et al., 2020; Myhr et al., 2020), positive rates of all subscales were further compared between males and females using Chi-square test; significance of trends were also tested in males and females separately.

## 3. Results

Demographic and mental health characteristics of students in each wave were shown in **Table 1**. There was no significant difference in sex ratio among the three waves (*χ*^2^ = 1.873, *p* = 0.392). Data on participants’ age was only available for the second wave and could not be compared. Cochran–Armitage trend tests showed that positive rates of the obsessive–compulsive tendencies (*p* < 0.001), interpersonal sensitivity (*p* = 0.015), depression (*p* < 0.001), anxiety (*p* = 0.001), academic stress (*p* < 0.001) and emotional disturbance (*p* = 0.006) problems were significantly increased during the period of 2016 to 2020, while no significant increasing or decreasing trends were found in positive rates of the paranoid ideation, hostility, maladaptation and psychological imbalance problems (all *p* > 0.05). The trends in positive rates of each mental health problem were also visualized in **Figure 1**.

**Table 1.**
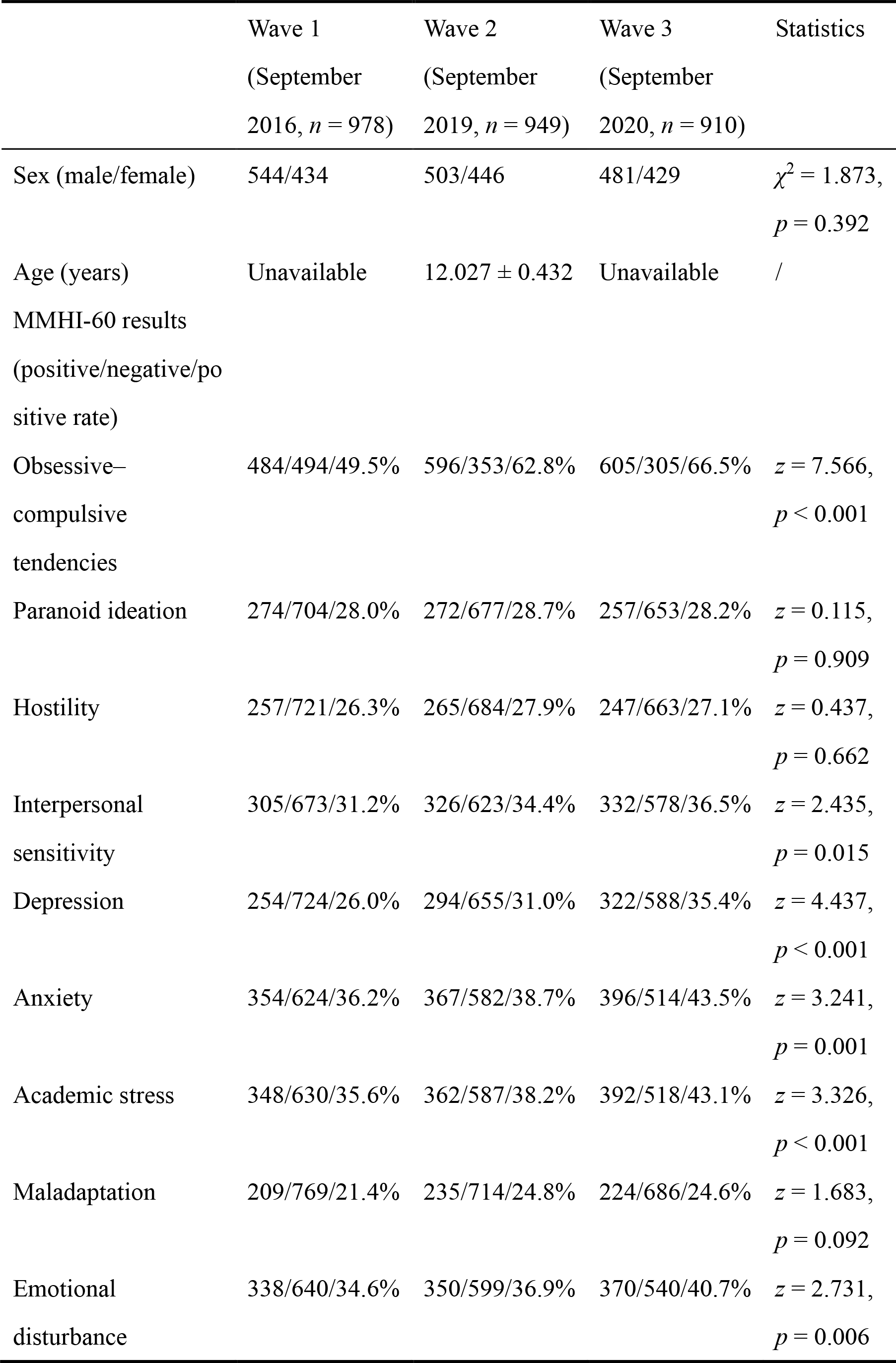
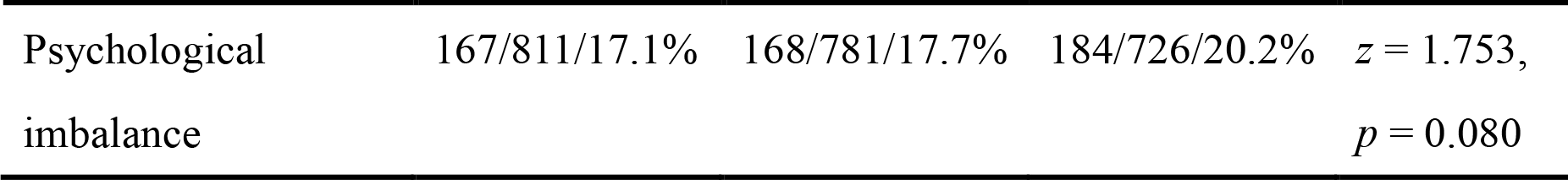
Sample characteristics of the participants surveyed in each wave.

**Figure 1.**
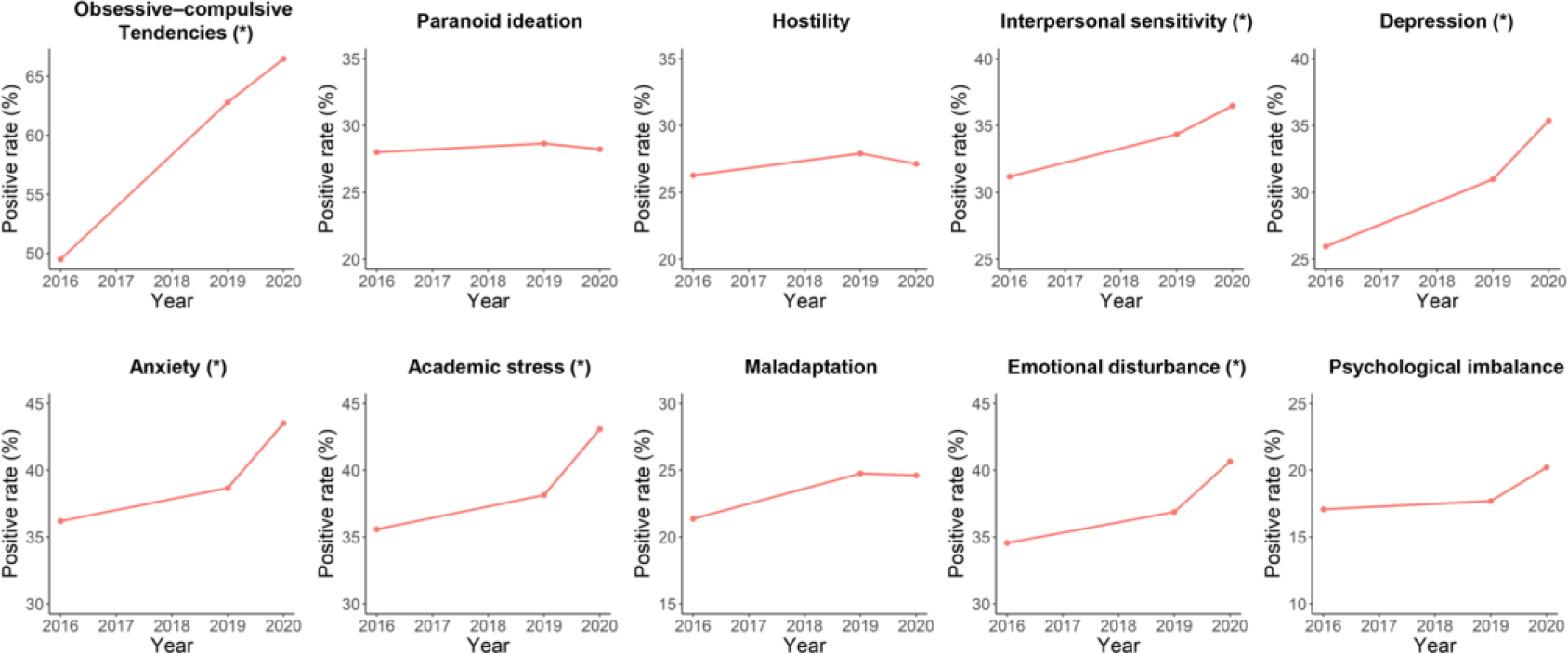
Trends in positive rates of each mental health problem in surveyed participants during the period of 2016-2020. The “*” indicates a significant increasing trend with *p* < 0.05.

The females had significantly higher positive rates of obsessive–compulsive tendencies, interpersonal sensitivity, depression, anxiety and academic stress, but a significantly lower positive rate of psychological imbalance than males (**Figure 2 and Supplementary Table 1**). During the period of 2016-2020, positive rate of obsessive– compulsive tendencies was significantly increased in both males and females, while positive rates of interpersonal sensitivity, depression, anxiety, academic stress, maladaptation, emotional disturbance and psychological imbalance were significantly increased in only females (**Figure 3 and Supplementary Tables 2-3**).

**Figure 2.**
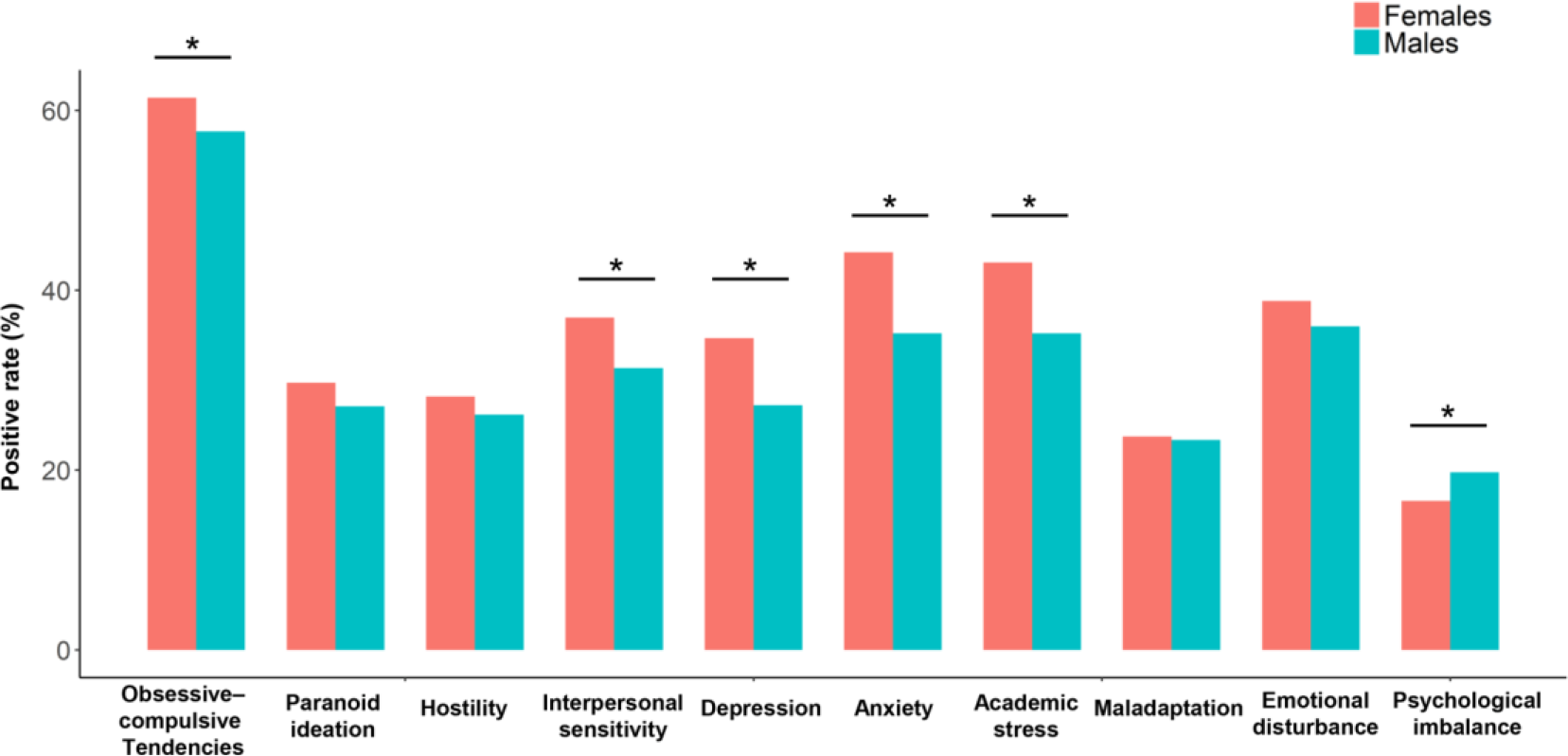
Comparisons on positive rates of each mental health problem between the female and male participants. The “*” indicates a significant sex difference with *p* < 0.05.

**Figure 3.**
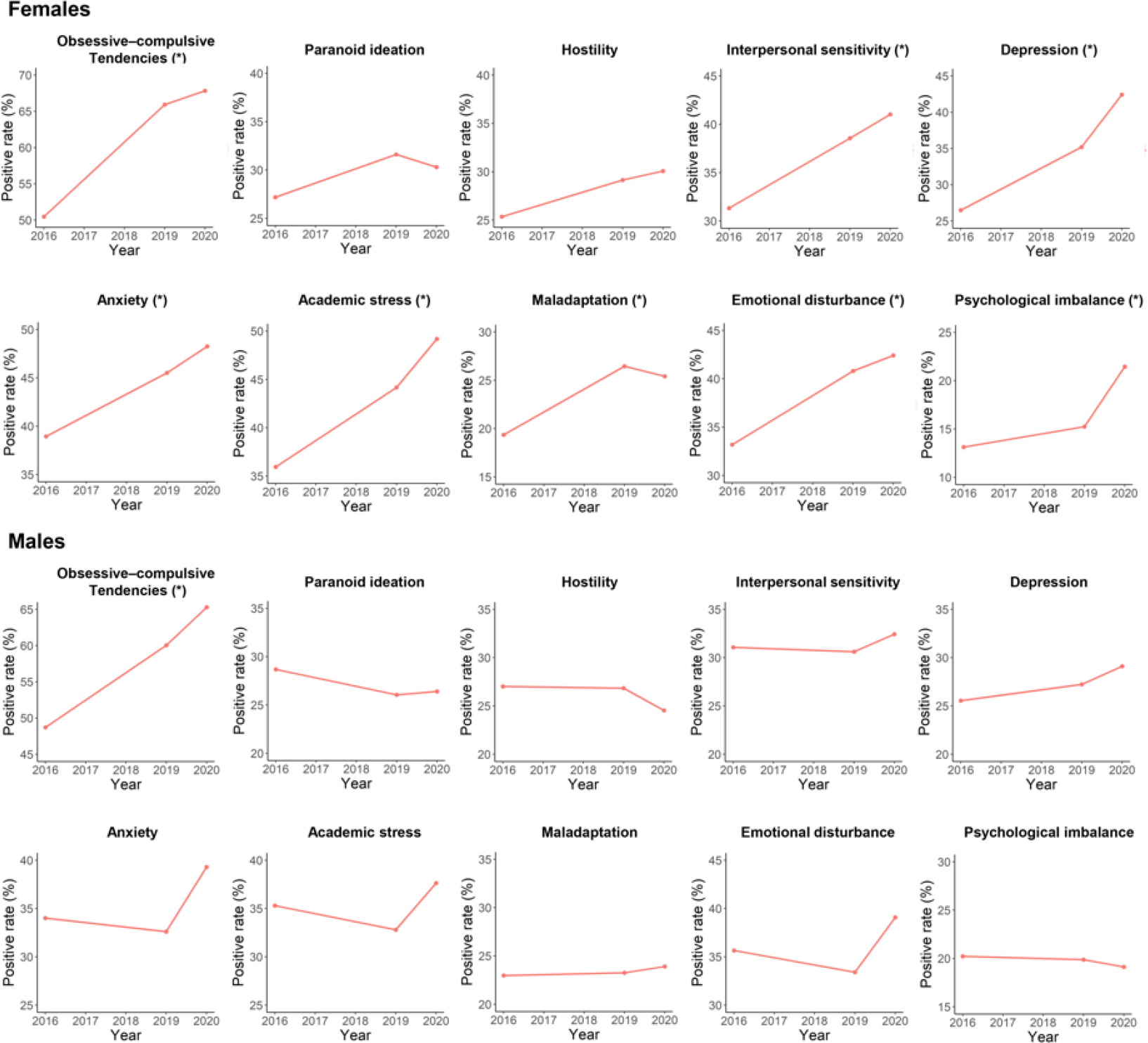
Trends in positive rates of each mental health problem during the period of 2016-2020 in surveyed females and males, respectively. The “*” indicates a significant increasing trend with *p* < 0.05.

## 4. Discussion

In this study, we performed a repeated cross-sectional analysis to explore the possible trends in mental health problems among Chinese adolescents over the years of 2016-2020. Compared with most published studies for similar purposes, we had advantages in methodologies (uniformity of surveyed populations) and data timeliness (until the most recent years).

Our results showed that positive rates of multiple mental health problems including obsessive–compulsive tendencies, interpersonal sensitivity, depression, anxiety, academic stress and emotional disturbance problems were all significantly increased during September 2016 to September 2020 in surveyed participants. Thus, as a supplement to previous studies analyzing data until 2018, our study suggests that mental health in Chinese adolescents could have been deteriorating in the most recent years. It is noteworthy that the COVID-19 outbreak in 2020, which has been widely reported to have negative mental effects on worldwide population (Ahmed et al., 2020; Rajkumar, 2020; Roy et al., 2020; Wu et al., 2021b). Therefore, the pandemic-related effects may play an important role in these trends besides other reported social and psychological factors (e.g., increasing divorce rate in China (Mo, 2017)).

Further analyses showed that positive rates of most of the above mental health problems were significantly higher in females than males and were significantly increased in only females during 2016-2020. Although being somewhat surprising, similar results have been reported among adolescents in other countries (Eyuboglu et al., 2021; Holik et al., 2021; Thorisdottir et al., 2021). It highlights the importance of focusing on girls since they may be more affected by mental problems.

Our study has several limitations. First, this survey was not performed in years of 2017 and 2018. Second, data on age was lacking for most participants. However, since children were required to enter school until 6 years old in China (Zhang et al., 2017), all participants should had a close age around 12 when they were surveyed. Third, the survey was performed in only a single school in Changsha city, and in only seventh-grade students in early adolescence. Thus, it should be cautious to interpret our results as trends in nationwide populations. Further studies conducted in a larger sample, in wider regions including rural areas, and in a wider range of age would be necessary to expand our knowledge in mental health among Chinese adolescents.

## 5. Conclusion

In this study, we found that positive rates of multiple mental health problems were significantly increased during 2016-2020 in a group of surveyed adolescents in Changsha city, China. Moreover, positive rates of most of these problems were found to be significantly higher in females than males, and significantly increased during 2016-2020 in only females. Such results highlight the importance of focusing on mental health problems among Chinese adolescents, especially among girls.

## Data Availability

All data produced in the present study are available upon reasonable request to the authors.

## Funding

This work was supported by the Natural Science Foundation of Hunan Province, China (2021JJ40851 to YL, 2020JJ5800 to XC) and the National Natural Science Foundation of China (82071506 to ZL).

## Acknowledgments

We would like to thank all the subjects who served as research participants.

## Declaration of competing interest

The authors declare no conflict of interest.

## Supplementary data

Supplementary material related to this article can be found in the online version.

## Notes

### Competing Interest Statement

The authors have declared no competing interest.

### Author Declarations

The Ethics Committee of the Second Xiangya Hospital of Central South University gave ethical approval for this work.

### Summary of Updates

Figures 1-3 updated

