## Supplementary Tables for "Increasing trends in mental health problems among Chinese young adolescents: results from repeated cross-sectional data in Changsha 2016-2020"

**Supplementary Table 1** Comparisons on positive rates of each mental health problem as measured by the MMHI-60 subscales between males and females.

| MMHI-60 subscale | Males (*n* = 1528) | Females (*n* = 1309) | Statistics |
|---|---|---|---|
| | positive/negative/positive rate | | |
| Obsessive–compulsive tendencies | 881/647/57.7% | 804/505/61.4% | $\chi^2 = 4.141$, $p = 0.041$ |
| Paranoid ideation | 414/1114/27.1% | 389/920/29.7% | $\chi^2 = 2.390$, $p = 0.122$ |
| Hostility | 400/1128/26.2% | 369/940/28.2% | $\chi^2 = 1.444$, $p = 0.229$ |
| Interpersonal sensitivity | 479/1049/31.4% | 484/825/37.0% | $\chi^2 = 9.955$, $p = 0.002$ |
| Depression | 416/1112/27.2% | 454/855/34.7% | $\chi^2 = 18.443$, $p < 0.001$ |
| Anxiety | 538/990/35.2% | 579/730/44.2% | $\chi^2 = 24.045$, $p < 0.001$ |
| Academic stress | 538/990/35.2% | 564/745/43.1% | $\chi^2 = 18.414$, $p < 0.001$ |
| Maladaptation | 357/1171/23.4% | 311/998/23.8% | $\chi^2 = 0.061$, $p = 0.805$ |
| Emotional disturbance | 550/978/36.0% | 508/801/38.8% | $\chi^2 = 2.386$, $p = 0.122$ |
| Psychological imbalance | 302/1226/19.8% | 217/1092/16.6% | $\chi^2 = 4.790$, $p = 0.029$ |

**Supplementary Table 2** Trends in each mental health problem in males.

| MMHI-60 subscale | Wave 1 (September 2016, *n* = 544) | Wave 2 (September 2019, *n* = 503) | Wave 3 (September 2020, *n* = 481) | Statistics |
|---|---|---|---|---|
| | positive/negative/positive rate | | | |
| Obsessive–compulsive tendencies | 265/279/48.7% | 302/201/60.0% | 314/167/65.3% | $z = 5.401$, $p < 0.001$ |
| Paranoid ideation | 156/388/28.7% | 131/372/26.0% | 127/354/26.4% | $z = -0.840$, $p = 0.401$ |
| Hostility | 147/397/27.0% | 135/368/26.8% | 118/363/24.5% | $z = -0.890$, $p = 0.374$ |
| Interpersonal sensitivity | 169/375/31.1% | 154/349/30.6% | 156/325/32.4% | $z = 0.445$, $p = 0.649$ |
| Depression | 139/405/25.6% | 137/366/27.2% | 140/341/29.1% | $z = 1.275$, $p = 0.202$ |
| Anxiety | 185/359/34.0% | 164/339/32.6% | 189/292/39.3% | $z = 1.714$, $p = 0.087$ |
| Academic stress | 192/352/35.3% | 165/338/32.8% | 181/300/37.6% | $z = 0.732$, $p = 0.464$ |
| Maladaptation | 125/419/23.0% | 117/386/23.3% | 115/366/23.9% | $z = 0.349$, $p = 0.727$ |
| Emotional disturbance | 194/350/35.7% | 168/335/33.4% | 188/293/39.1% | $z = 1.087$, $p = 0.278$ |
| Psychological imbalance | 110/434/20.2% | 100/403/19.9% | 92/389/19.1% | $z = -0.436$, $p = 0.663$ |

**Supplementary Table 3** Trends in each mental health problem in females.

| MMHI-60 subscale | Wave 1 (September 2016, *n* = 544) | Wave 2 (September 2019, *n* = 503) | Wave 3 (September 2020, *n* = 481) | Statistics |
|---|---|---|---|---|
| | positive/negative/positive rate | | | |
| Obsessive–compulsive tendencies | 219/215/50.5% | 294/152/65.9% | 291/138/67.8% | $z = 5.250$, $p < 0.001$ |
| Paranoid ideation | 118/316/27.2% | 141/305/31.6% | 130/299/30.3% | $z = 1.004$, $p = 0.315$ |
| Hostility | 110/324/25.4% | 130/316/29.2% | 129/300/30.1% | $z = 1.544$, $p = 0.123$ |
| Interpersonal sensitivity | 136/298/31.3% | 172/274/38.6% | 176/253/41.0% | $z = 2.951$, $p = 0.003$ |
| Depression | 115/319/26.5% | 157/289/35.2% | 182/247/42.4% | $z = 4.916$, $p < 0.001$ |
| Anxiety | 169/265/38.9% | 203/243/45.5% | 207/222/48.3% | $z = 2.756$, $p = 0.006$ |
| Academic stress | 156/278/35.9% | 197/249/44.2% | 211/218/49.2% | $z = 3.929$, $p < 0.001$ |
| Maladaptation | 84/350/19.4% | 118/328/26.5% | 109/320/25.4% | $z = 2.095$, $p = 0.036$ |
| Emotional disturbance | 144/290/33.2% | 182/264/40.8% | 182/247/42.4% | $z = 2.790$, $p = 0.005$ |
| Psychological imbalance | 57/377/13.1% | 68/378/15.3% | 92/337/21.5% | $z = 3.280$, $p = 0.001$ |
